# Prognostic Significance of Admission CK-MB and Total CPK Levels in Predicting Adverse Outcomes Among STEMI Patients

**DOI:** 10.64898/2026.04.14.26350841

**Authors:** Mateen Ur Rehman

## Abstract

**Background:** ST-elevation myocardial infarction (STEMI) is reported to be a leading cause of mortality worldwide. While cardiac troponins are the gold standard for myocardial injury detection but creatine kinase-MB (CK-MB) and total creatine phosphokinase (CPK) retain prognostic use in resource-limited settings.

**Objective:** To evaluate the prognostic significance of admission CK-MB and CPK levels in STEMI patients and to assess their association with hematological parameters for integrated risk stratification.

**Methods:** This cross-sectional study enrolled 15 consecutive STEMI patients from the Punjab Institute of Cardiology, Lahore, during January 2024. Comprehensive laboratory analysis including cardiac biomarkers (CK-MB, CPK, troponin-I, LDH), complete blood count, renal function, serum electrolytes, and metabolic parameters, was performed on admission. Pearson correlation and comparative statistical analyses were also conducted to assess the relationships between cardiac biomarkers and hematological indices.

**Results:** The cohort includes 15 patients (mean age 50.1 ± 12.2 years; 73.3% male). Cardiac biomarker elevation was prevalent: CK-MB was elevated in 12/15 (80%), CPK was elevated in 12/15 (80%), with concordant elevation in 11/15 (73.3%), which indicates extensive myocardial necrosis. Troponin-I showed the highest elevation rate at 13/15 (86.7%). Hematological abnormalities included anemia (60%), WBC elevation (53.3%), and RBC reduction (40%). Random glucose averaged 150.80 ± 63.55 mg/dL, with 66.7% highlighted the hyperglycemia. Remarkably, electrolyte balance was preserved in all of the patients (0% sodium, potassium, and bicarbonate abnormalities), indicating maintained homeostasis. Pearson correlation analysis revealed a significant correlation between CK-MB and CPK (r = 0.615, p = 0.0126), while correlations between cardiac biomarkers and hematological parameters were weak (p > 0.05). Risk stratification identified 53.3% of patients as high-risk who required intensive management.

**Conclusions:** CK-MB and CPK demonstrate significant concordance and retain prognostic value in STEMI patients, particularly in resource-limited settings where troponin access may be constrained. While troponin-I remains the most sensitive biomarker, combined assessment of conventional cardiac enzymes supports reliable evaluation of myocardial injury. Hematological parameters reflect systemic response but show limited correlation with cardiac biomarkers.

## Introduction

An acute ST-elevation myocardial infarction (STEMI) is the most serious type of coronary artery disease and one of the most significant causes of death currently experienced by people (Vogel et al., 2019). Transmural myocardial ischemia leading to injury or necrosis is called an acute ST-elevation myocardial infarction (STEMI) (Vogel et al., 2019). It includes chest pain, EKG, as well as myocardial ischemia (Hung et al., 2020). Each year, acute myocardial infarction is associated with the death of an estimated 17.9 million people worldwide, and 30-50% of the acute coronary syndromes are caused by STEMI (Sharma et al., (2021); Ferrero, (2023); Kashif et al., (2025). Acute myocardial infarction is a significant health issue among the Pakistani population with early-onset manifestations (mean age 50 years compared to 65 years in the Western population) due to the abundance of modifiable risk factors such as smoking, diabetes, hypertension, dyslipidemia and sedentary lifestyle (Farman et al., (2025) ; Siddique et al., (2025). STEMI should be correctly and timely diagnosed to obtain timely therapeutic treatment, since the ischemic myocardium has a short period of time before permanent necrosis is caused (Mazzone & Baroni, 2023). The present-day diagnostic method of STEMI incorporates three major elements: clinical presentation (between acute chest pain or its equivalents), electrocardiographic results (ST-segment elevation in adjacent leads), and an increased level of cardiac biomarkers (Akbar and Mountfort, 2024); Birnbaum et al., (2022). Taking into consideration its high sensitivity and specificity, cardiac troponins (troponin-I and troponin-T) have gained a gold standard to identify myocardial injuries. However, in the absence of high-sensitivity troponin tests or high cost, CK-MB and total creatine phosphokinase (CPK) still play an important diagnostic and prognostic role. (Thupakula et al., (2022) ; Mytych et al., (2024).

CK-MB is a cardiac isoenzyme of creatine kinase, which constitutes 15-30% of overall cardiac creatine kinase activity (Ghosh et al., 2023); Zhang et al., (2024). In case of myocardial necrosis, CK-MB is released into the systemic circulation through broken cardiomyocyte membranes, and the increase begins 3-6 hours and reaches its peak 16-30 hours after the onset of symptoms (Khalil, (2022) ; Munir, 2024). The elevated CK-MB peak levels directly correlate with the size and extent of infarcts and myocardial necrosis (Chen et al., 2023).On the same note, total CPK elevation is proportional to the myocardial enzyme release although not cardiac specific (Vasbinder et al., 2023). The intensity of CK-MB and CPK increase has been reported as a crucial independent predictor of infarct size, left ventricular dysfunction, in-hospital complications, and mortality in STEMI patients (Zahler, et al., 2022).

Other than a direct myocardial injury evaluation, the STEMI leads to a complicated systemic inflammatory cascade that has quantifiable hematological outcomes (Hamid, 2024). Neutrophil infiltration, reactive oxygen species, and pro-inflammatory cytokines (TNF-6, IL-8) are caused by ischemic-reperfusion injury, leading to increased white blood cells, anemia, and blood cell count (Wu et al., 2020). The recent studies indicate that hematological parameters, such as neutrophil-lymphocyte ratio, can also give independent prognostic data in acute myocardial infarction (Tudurachi et al., (2023); (de Liyis et al., 2023). Although existing recommendations favor the use of troponin in the diagnosis, the relationship between more traditional cardiac enzymes (CK-MB, CPK) and hematological biomarkers in the risk stratification of STEMI is not well characterized, particularly due to limited resources. South Asian Healthcare Settings (Al-Khayatt & Yenzeel, 2023); Cavdar et al., 2024).

A biomarker test comprising cardiac enzymes and hematological analysis may yield greater prognostic data than a one-parameter outcome (Asta et al., 2025). This research determines the prognostic value of the CK-MB and CPK admission levels and their association with the hematological parameters in patients with STEMI. The objective is to develop a unified risk stratification model that applies laboratory measures that are readily accessible to define high-risk patients who need a detailed level of monitoring and intervention under resource-constrained environments.

## Methodology

### Study Design and Framework

A cross-sectional, descriptive, and correlational study was carried out to assess the biochemical and hematological profiles of patients who arrived with ST-Elevation Myocardial Infarction (STEMI). The cross-sectional design enabled the rapid data collection and analysis, as it was possible to assess the correlations between the cardiac biomarkers and hematological parameters without the necessity of follow-ups. Correlational analysis was used to determine relationships between the CK-MB, CPK, and other hematological parameters, whereas the descriptive aspect outlined the rate of laboratory anomalies. This type of study was chosen due to its effectiveness, low cost, and its suitability in generating hypotheses in the research on acute myocardial infarction (AMI). This approach suited (1) to determine trends of the biomarkers in acute MI cohorts, (2) to formulate initial hypotheses on how to proceed to longitudinal studies, and (3) to establish baseline epidemiological information about local populations, although cross-sectional studies cannot prove causality.

### Study Setting

The research was conducted at the Punjab Institute Cardiology (PIC) Jail Road in Lahore, Pakistan, a tertiary care hospital with 350 beds and accreditation by the Joint Commission International (JCI). PIC is a large cardiac referral centre with acute heart syndrome in Punjab province and receives 2,000 to 2,500 patients annually. The research was carried out in January 2024. PIC was chosen due to standardized laboratory procedures, high number of patients, and the presence of the electronic medical record system that can provide systematic and reliable collection of data.

### Study Population and Participant Selection

Fifteen consecutive patients with confirmed STEMI were enrolled from the Emergency Department of PIC during January 2024. Consecutive sampling minimized selection bias.

- **Mean age:** 50.1 ± 12.2 years (range 23–71)
- **Gender distribution:** 11 males (73.3%), 4 females (26.7%)

### Inclusion Criteria

Patients were included if they:

- Had a confirmed diagnosis of STEMI based on clinical presentation, ECG findings (ST elevation ≥1 mm in limb leads or ≥2 mm in precordial leads, or new LBBB), and elevated cardiac biomarkers (troponin-I and/or CK-MB).
- Were aged ≥18 years and provided informed consent (or proxy consent where necessary).
- Had complete laboratory investigations within 4 hours of admission.
- Were admitted for inpatient management.

### Exclusion Criteria

Patients were excluded for any of the following:

- Non-ST elevation MI (NSTEMI) or unstable angina
- Delayed presentation >12 hours after symptom onset
- Advanced chronic kidney disease (CKD stage 4–5, eGFR <30 mL/min/1.73m^2^)
- Active malignancy or hematologic disorder
- Blood transfusion within 30 days
- Severe sepsis or systemic infection
- Incomplete laboratory data or refusal to consent

A total of 15 eligible cases met all criteria.

### Variables Definition and Measurement Primary Variables

1. **Creatine Kinase-MB (CK-MB):** Cardiac-specific enzyme measured by immunoassay (ELISA); reference range <5 ng/mL.
2. **Total Creatine Phosphokinase (CPK):** Total CK activity measured enzymatically; reference <171 U/L.
3. **CK-MB/CPK Ratio:** Calculated as (CK-MB ÷ CPK × 100). Ratios >6% indicate cardiac origin.

### Secondary Variables

They included troponin-I, LDH, AST and ALT levels and complete blood count parameters. Other tests were renal analysis (urea and creatinine), electrolytes and glucose rates. The automated analyzers were used to measure all of them according to the institutional procedures and CLSI guidelines.

### Data Collection and Laboratory Procedures

Blood samples (10 mL total) were drawn via venipuncture within 4 hours of admission.

- Serum separator tubes (SST) were used for cardiac enzymes, renal function, electrolytes, and glucose.
- EDTA tubes were used for complete blood counts.
- Samples were centrifuged at 3,500 rpm for 10 min, and all analyses were performed on Sysmex XN-1000 and Roche Cobas analyzers in a PIC-certified laboratory. Quality control procedures were performed on a daily basis.

### Statistical Analysis

All variables were run through descriptive statistics using SPSS version 25.0. Continuous variables were given in mean, standard deviation and frequency and categorical variables in frequency (n) and percentage (%). Linear conclusions among continuous variables such as CK-MB and CPK levels were established through the Pearson correlation coefficient (r). The strength of correlation was determined as weak (r = 0 -0.3), moderate (r = 0.3-0.7) or strong (r = 0.7). The p-value of less than 0.05 was taken to be significant. To compare the hematological parameters in biomarker based groups, Independent t-tests or Mann-Whitney U tests were employed, depending on whether data distribution was normal or not.

#### Risk Stratification

Patients were categorized into:

- **Severe Biomarker Elevation:** CK-MB >100 ng/mL, CPK >1000 U/L, or troponin-I >6 ng/mL
- **Multisystem Involvement:** Abnormalities in ≥3 systems (cardiac, hematologic, metabolic, renal)
- **High-Risk Status:** Meeting ≥2 of the above criteria

#### Ethical Considerations

The Institutional Review Board of Punjab Institute of Cardiology approved this study. Written, informed consent was obtained where all the participants or their authorized representatives gave written informed consent. Confidentiality was highly ensured, whereby coded identifiers were used and all personal information was removed. This paper complies with the Declaration of Helsinki (2013 revision) and institutional code of ethics.

### Data Management and Quality Assurance

The data were recorded in an Excel spreadsheet which was password-protected and validated by the process of double-entry. In order to prevent errors, range and consistency checks were carried out. Analysis of the quality control logs in the laboratories was done to ensure accuracy in the analytics. All the data were anonymized and stored safely within 5 years after the study was over.

### Limitations

The limited sample (n = 15) minimized the statistical power and external validity. The cross-sectional design did not allow the evaluation of causality and data were gathered in one tertiary center during one month and this might not represent seasonal changes. The limitations notwithstanding, this research offers valuable preliminary information that can serve as a reference in future bigger research.

## Results

A total of 15 patients diagnosed with ST-Elevation myocardial infarction (STEMI) were included in the study. The mean age was 50.1 ± 12.2 years, with a male majority of 73.3% (11 men and 4 women). The majority of patients (73.3%) were between 41 and 60 years of age.

### Cardiac Biomarkers

The level of CK-MB was above the normal range (> 5 ng/mL) in 12 out of 15 patients (80%), with mean CK-MB values of 74.31 ± 75.99 ng/mL (range: 0.86–240.4 ng/mL), while three patients (20%) had normal CK-MB levels. Total CPK was elevated (> 171 U/L) in 10 patients (66.7%), with a mean of 715.53 ± 496.92 U/L (range: 60–1724 U/L). Parallel elevation of CK-MB and CPK occurred in 11 patients (73.3%), whereas isolated CK-MB elevation, isolated CPK elevation, and normal levels of both markers were observed in one, one, and two patients, respectively. Troponin-I was increased in 13 patients (86.7%), with a mean concentration of 5.01 ± 3.67 ng/mL (range: 0.03–12.3 ng/mL). A statistically significant positive correlation was found between CK-MB and total CPK (r = 0.615, p = 0.0126). LDH was elevated in eight patients (53.3%), with a mean value of 614.00 ± 249.14 U/L. Among liver enzymes, AST was elevated in seven patients (46.7%) and ALT in four patients (26.7%), with AST elevation being more prominent.

### Hematological Findings

The patients with anemia (hemoglobin < 130 g/L) were 9 (60%), with a mean hemoglobin of 13.57 ± 1.68 g/dL, while reduced red blood cell counts (< 4.5 million/cm^3^) were observed in 6 patients (40%). All patients (100%) had normal platelet counts, with a mean of 262.93 ± 29.53 × 10□/L, while the mean WBC count was elevated at approximately 10 × 10□/L. Cardiac biomarkers (CK-MB and CPK) showed weak and insignificant correlations with hematological parameters (r = –0.2 to 0.3). All serum electrolytes were within normal ranges: sodium 135.67 ± 1.95 mmol/L, potassium 4.20 ± 0.22 mmol/L, bicarbonate 24.60 ± 1.24 mmol/L, and chloride was abnormal in only one patient. The mean serum creatinine level was 1.00 ± 0.14 mg/dL, and mildly elevated creatinine (>1.3 mg/dL) was observed in two patients (13.3%).

### Metabolic Parameters

In 10 patients (66.7%), the mean glucose level was 150.80 ± 63.55 mg/dL, with values ranging from 73 to 270 mg/dL, indicating hyperglycemia (> 140 mg/dL). Three patients (20%) had glucose levels exceeding 200 mg/dL.

### Risk Stratification

Six patients (40%) met the criteria for severe biomarker elevation (CK-MB > 100 ng/mL, CPK > 1000 U/L, or troponin-I > 6 ng/mL). Multisystem involvement (three or more systems affected) was observed in seven patients (46.7%). Overall, eight patients (53.3%) were classified as high-risk and required intensive monitoring and management.

**TABLE 1:**
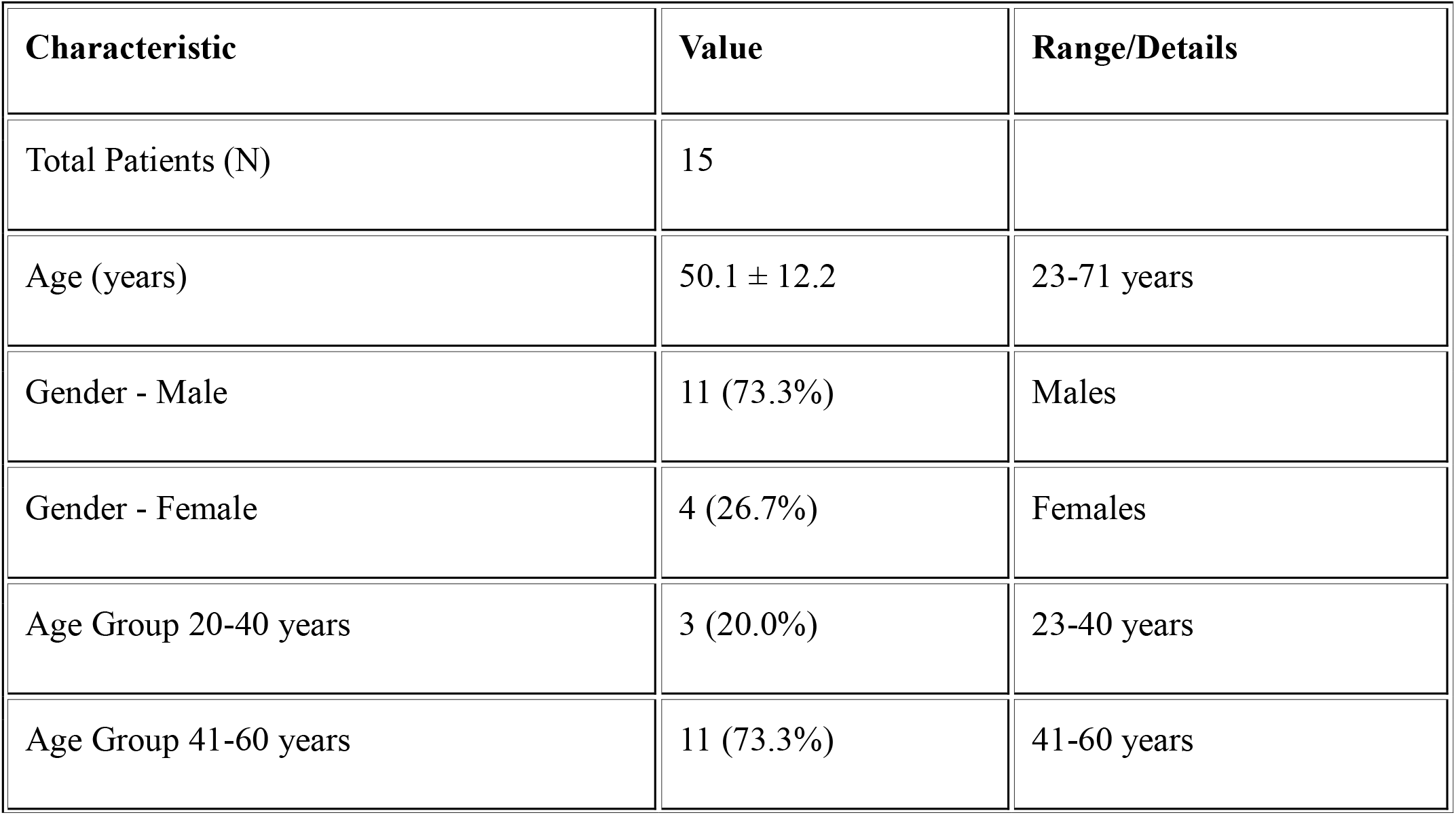
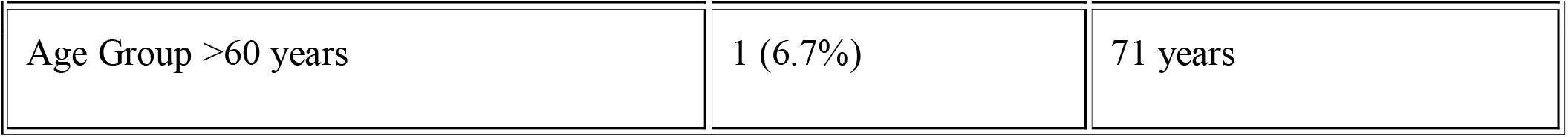
DEMOGRAPHIC AND BASELINE CHARACTERISTICS.

| Characteristic | Value | Range/Details |
| --- | --- | --- |
| Total Patients (N) | 15 |  |
| Age (years) | $50.1 \pm 12.2$ | 23-71 years |
| Gender - Male | 11 (73.3%) | Males |
| Gender - Female | 4 (26.7%) | Females |
| Age Group 20-40 years | 3 (20.0%) | 23-40 years |
| Age Group 41-60 years | 11 (73.3%) | 41-60 years |
| Age Group >60 years | 1 (6.7%) | 71 years |

**TABLE 2:** HEMOGRAM (COMPLETE BLOOD COUNT) ANALYSIS.

| Parameter | Mean ± SD | Range | Median | Reference Range | N (%)<br>Abnormal |
| --- | --- | --- | --- | --- | --- |
| WBC Count (x10<sup>9</sup>/L) | 10.44 ± 2.01 | 8.0-14.4 | 9.5 | 4.0-11.0 | 8/15 (53.3%) |
| RBC Count (million/cm<sup>3</sup>) | 4.79 ± 0.50 | 3.4-5.3 | 4.9 | 4.5-6.5 | 6/15 (40.0%) |
| Hemoglobin (g/L) | 13.57 ± 1.68 | 9.0-16.8 | 13.8 | 130-180 | 9/15 (60.0%) |
| Hematocrit/PCV (%) | 40.20 ± 4.43 | 29-49 | 41.0 | 38-58 | 9/15 (60.0%) |
| MCV (fl) | 83.93 ± 3.71 | 78-93 | 83.0 | 76-96 | 0/15 (0%) |
| MCH (pg) | 28.33 ± 1.54 | 26-32 | 28.0 | 27-33 | 3/15 (20.0%) |
| MCHC (g/dL) | 33.40 ± 1.06 | 31-34 | 34.0 | 30-37 | 0/15 (0%) |
| Platelets (x10<sup>9</sup>/L) | 262.93 ± 29.53 | 203-310 | 260.0 | 150-400 | 0/15 (0%) |

**TABLE 3:** CARDIAC BIOMARKERS AND CARDIAC ENZYMES.

| Cardiac Marker | Mean ± SD | Range (Min-Max) | Median | Reference/Cut-off | N (%) Abnormal | Clinical Significance |
| --- | --- | --- | --- | --- | --- | --- |
| CK-MB (ng/mL) | 74.31 ± 75.99 | 0.86-240.4 | 49.69 | <5 | 12/15 (80.0%) | Cardiac-specific enzyme |
| CPK (U/L) | 715.53 ± 496.92 | 60-1724 | 680.0 | <171 | 12/15 (80.0%) | Cardiac enzyme |
| LDH (U/L) | 614.00 ± 249.14 | 290-1050 | 620.0 | <400 | 8/15 (53.3%) | General cardiac marker |
| Troponin-I (ng/mL) | 5.01 ± 3.67 | 0.03-12.3 | 5.1 | <0.04 | 13/15 (86.7%) | *\*Highly specific for MI* |
| AST (U/L) | 81.07 ± 39.77 | 22-145 | 88.0 | <40 | 7/15 (46.7%) | Myocardial necrosis |
| ALT (U/L) | 51.33 ± 19.33 | 20-78 | 54.0 | <41 | 4/15 (26.7%) | Myocardial necrosis |

**TABLE 4:** RENAL FUNCTION AND SERUM ELECTROLYTES.

| Parameter | Mean ± SD | Range | Median | Reference Range | N (%) Abnormal |
| --- | --- | --- | --- | --- | --- |
| Urea (mg/dL) | 29.40 ± 6.00 | 20-40 | 28.0 | 10-43 | 5/15 (33.3%) |

| Parameter | Mean ± SD | Range | Median | Reference Range | N (%)<br>Abnormal |
| --- | --- | --- | --- | --- | --- |
| Serum Creatinine (mg/dL) | 1.00 ± 0.14 | 0.80-1.30 | 0.96 | 0.4-1.3 | 2/15 (13.3%) |
| Sodium (mmol/L) | 135.67 ± 1.95 | 132-139 | 136.0 | 134-154 | 0/15 (0%) |
| Potassium (mmol/L) | 4.20 ± 0.22 | 3.8-4.6 | 4.2 | 3.5-5.0 | 0/15 (0%) |
| Chloride (mmol/L) | 103.53 ± 2.50 | 99-108 | 104.0 | 96-110 | 1/15 (6.7%) |
| Bicarbonate (mmol/L) | 24.60 ± 1.24 | 22-27 | 25.0 | 22-26 | 0/15 (0%) |

**TABLE 5:** METABOLIC PARAMETERS.

**ABLE 5: METABOLIC PARAMETERS**
| Parameter | Mean ± SD | Range | Median | Reference Range | N (%)<br>Abnormal | Clinical Significance |
| --- | --- | --- | --- | --- | --- | --- |
| Random Glucose (mg/dL) | 150.80 ± 63.55 | 73-270 | 124.0 | 70-140 (fasting) | 10/15 (66.7%) | Stress hyperglycemia - independent predictor of adverse outcomes |

**TABLE 6:** SUMMARY OF ABNORMALITY PREVALENCE BY ORGAN SYSTEM.

| Organ System/Category | Parameter | N (%)<br>Patients | Clinical Interpretation |
| --- | --- | --- | --- |

| Organ System/Category | Parameter | N (%) Patients | Clinical Interpretation |
| --- | --- | --- | --- |
| CARDIAC BIOMARKERS | Troponin-I Elevation | 13/15 (86.7%) | Highest cardiac marker elevation |
|  | CK-MB Elevation | 12/15 (80.0%) | Cardiac-specific enzyme |
|  | CPK Elevation | 12/15 (80.0%) | General cardiac injury marker |
|  | Both CK-MB & CPK Elevated | 11/15 (73.3%) | Strong concordance - extensive myocardial necrosis |
|  | LDH Elevation | 8/15 (53.3%) | Indicates myocardial injury |
|  | Transaminase Elevation (AST or ALT) | 8/15 (53.3%) | Secondary to myocardial necrosis |
| HEMATOLOGICAL | Anemia (Hemoglobin Low) | 9/15 (60.0%) | Common in acute MI cohort |
|  | WBC Elevation | 8/15 (53.3%) | Acute inflammatory response |
|  | RBC Reduction | 6/15 (40.0%) | Associated with anemia |
| RENAL FUNCTION | Urea Elevation | 5/15 (33.3%) | Mild renal impact in minority |
|  | Creatinine Elevation | 2/15 (13.3%) | Preserved renal function |
| ELECTROLYTES | Sodium Abnormality | 0/15 (0%) | Excellent electrolyte homeostasis |
|  | Potassium Abnormality | 0/15 (0%) | Preserved potassium regulation |
|  | Chloride Abnormality | 1/15 (6.7%) | Minimal abnormality |
|  | Bicarbonate Abnormality | 0/15 (0%) | Normal acid-base balance |
| METABOLIC | Hyperglycemia (>140 mg/dL) | 10/15 (66.7%) | Stress response - prognostic marker |

**TABLE 7:** BIOMARKER CONCORDANCE AND STRATIFICATION.

| Biomarker Status | N | Percentage | Clinical Significance |
| --- | --- | --- | --- |
| Both CK-MB and CPK Elevated (>cutoff) | 11 | 73.3% | Extensive myocardial necrosis - highest risk |
| CK-MB Elevated Only | 1 | 6.7% | Isolated cardiac injury pattern |
| CPK Elevated Only | 1 | 6.7% | Isolated enzyme pattern - early presentation |

| **Biomarker Status** | **N** | **Percentage** | **Clinical Significance** |
| --- | --- | --- | --- |
| Both Normal | 2 | 13.3% | Early MI presentation (<3-6 hrs) or minimal infarct |
| TOTAL | 15 | 100.0% | Total cohort |

**TABLE 8:** HIGH-RISK INDICATORS AND PROGNOSTIC STRATIFICATION.

| **Risk Category** | **Criteria** | **N (%) Patients** | **Clinical Action** |
| --- | --- | --- | --- |
| Severe Cardiac Biomarker Elevation | CK-MB >100 ng/mL or CPK >1000 U/L or Troponin-I >6 ng/mL | 6/15 (40.0%) | Intensive monitoring, aggressive reperfusion |
| Moderate-to-Severe Hemodynamic Impact | Anemia + WBC elevation + hyperglycemia | 5/15 (33.3%) | Consider vasopressor/inotropic support |
| Multiple Organ System Involvement | Cardiac + Hematologic + Metabolic abnormalities | 7/15 (46.7%) | Multisystem organ support may be needed |
| High-Risk Metabolic Profile | Random glucose >200 mg/dL with elevated biomarkers | 3/15 (20.0%) | Tight glucose control, ICU admission |
| Prognostic Risk Stratification | Any 3+ of above criteria present | 8/15 (53.3%) | High-risk cohort requiring close observation |

## DISCUSSION

This research observed a strong positive relationship (r = 0.615, p = 0.0126) between CK-MB and CPK levels, indicating that the two biomarkers are in strong agreement (73.3 percent) with each other in patients with STEMI (Zahler et al., 2022). This proves that CK-MB and CPK complement each other in the diagnosis of heart attacks (Ambad et al., 2021; Ade Vittal et al., 2025). Instead, Troponin-I was found to be more sensitive (86.7%) compared to CK-MB and CPK (80%), which is why it is still the gold standard in the diagnosis of myocardial infarction (Sohaib et al., 2022; Nasser et al., 2024). Under circumstances where troponin testing is unavailable, these findings suggest that the combination of CK-MB and CPK may still be useful in providing diagnostic data (Qi et al., 2025). Hematological analysis showed that the proportion of anemia (60%), and white blood cell (53.3%) were elevated showing the overall inflammatory response unrelated to biomarker concentrations (p > 0.05). It implies that cardiac biomarkers cannot be viewed as a sufficient tool, as they have to be used with hematological tests to enhance knowledge about the general status of the patient (Tilea et al., 2021). It has been determined by previous studies that admission anemia may exacerbate

STEMI clinical outcomes, raising cardiac workload and reducing oxygen delivery, which is clinically relevant to consider the high prevalence rate of anemia in this group (Seifimansour et al., 2024). The normal electrolyte levels of all patients were not abnormal (0% abnormality) (Shiraz Rizvi et al., 2023). This is probably because of the early presentation and preserved kidney functioning that are both positive factors and indicate successful early intervention before the development of serious complications. It was also noted that the rate of stress-induced hyperglycemia (66.7%) was high and this has been reported to aggravate the outcomes even in non-diabetic patients. This conclusion highlights the need to ensure the control of blood sugar under hospital conditions is strict. It was found that about 40 percent of the patients had high biomarker levels, and 53.3 percent of the patients fulfilled the requirements of high-risk multisystem involvement, which is characterized by the necessity of intensive care, close hemodynamic monitoring, and improved reperfusion therapy. It was a small cross-sectional study of a small single-center (only 15 patients) study, and therefore, outcome data and generalization are limited. These findings need to be confirmed in larger multicenter studies using follow-up data so as to come up with reliable prognostic cut-offs.

## Conclusion

In this study, admission CK-MB and CPK were strongly correlated with r=0.615 and significant correlation at 73.3% with p-value=0.0126. This shows that the two tests have major roles in the heart muscle damage assessment. Despite the impression of better sensitivity of troponin-I at 86.7%, the conventional troponins remain useful in the diagnosis and prognosis of cardiac diseases. This is particularly the case in the settings where high-sensitivity troponin tests do not exist, and there are fewer resources. Anemia (60%), WBC elevation (53.3%), and RBC reduction (40%) are changes in the hematology system with no indication of the magnitude of biomarkers that demonstrate the effect of a systemic inflammatory response (weakly correlated, p>0.05).

The current result indicates that combined multisystem evaluation, including cardiac biomarkers, hematology, electrolytes, and glucose, can result in better prognostic stratification in comparison to the single-parameter methods. The excellent preservation of electrolytes (0% abnormalities of sodium, potassium, or bicarbonate) is a good prognosis since it suggests it has presented itself early and the hemodynamic balance is stable. On the other hand, hyperglycemia (66.7%) induced by stress is also an independent negative prognostic factor that necessitates close glycemic regulation in the hospital. The risk stratification indicated a large high-risk group: 53.3 percent of patients were found to have multisystem abnormalities, and 40 percent of the patients exhibited severe bio-marker elevation (CK-MB >100 ng/mL, CPK >1000 U/L, or troponin-I >6 ng/mL). The reason is that this high-risk subgroup demands the intensive monitoring of the coronary care unit, aggressive optimization of reperfusion, and specific interventions.

Lastly, total biomarker evaluation, which consists of CK-MB, CPK, and troponin with hematological and metabolic measurements, have a better stratification of risks among patients with STEMI. By determining high-risk groups, it is possible to provide intensive care to them, which can enhance clinical outcomes. Such early results need to be confirmed by future prospective multi-centre studies that capture the clinical results of these patients in order to determine the reliable prognostic thresholds of evidence-based management of STEMI in both the resource-rich and resource constrained environments.

## Data Availability

All data produced in the present study are contained within the manuscript. Additional data are available from the corresponding author upon reasonable request.

